# Biomarkers of insulin resistance and their performance as predictors of treatment response in overweight adults

**DOI:** 10.1101/2024.10.06.24314962

**Authors:** RJ Brogan, O Rooyackers, BE Phillips, B Twelkmeyer, LM Ross, PJ Atherton, WE Kraus, JA Timmons, IJ Gallagher

**Author notes:** Senior Authors (JA Timmons, IJ Gallagher).

## Abstract

Insulin Resistance (IR) contributes to the pathogenesis of type 2 diabetes mellitus (T2DM), and is a risk factor for cardiovascular and neurodegenerative diseases. Amino acid and lipid metabolomic biomarkers associate with future T2DM risk in epidemiological cohorts. Whether these biomarkers can accurately detect changes in IR status following treatment is unclear. Herein we evaluated the performance of clinical and metabolomic biomarkers to predict altered IR following lifestyle-based interventions. First, we evaluated the performance of two distinct insulin assay types (high-sensitivity ELISA and Immunoassay) and built cross-sectional clinical and metabolomic IR diagnostic models. These were utilised to stratify IR status in pre-intervention fasting samples, from three independent cohorts (META-PREDICT (MP, n=179), STRRIDE-AT/RT (S-2, n=116) and STRRIDE-PD (S-PD, n=149)). Linear and Bayesian projective prediction strategies were used to evaluate biomarkers for fasting insulin and HOMA2-IR and change in fasting insulin with treatment. Both insulin assays accurately quantified international standard insulin (R^2^>0.99), yet agreement for fasting insulin was less congruent (R^2^=0.65). A mean treatment effect on fasting insulin was only detectable using an ELISA. Clinical-metabolomic models were statistically related to fasting insulin (R^2^ 0.33-0.39) with modest capacity to classify IR at a clinically relevant HOMA2-IR threshold. Furthermore, no model predicted treatment responses in any cohort. Thus, we demonstrate that the choice of insulin assay is critical when quantifying the influence of treatment on fasting insulin, while none of the clinical-metabolomic biomarkers, established in cross-sectional data, are suitable for monitoring longitudinally changes in IR status.

## Introduction

Chronic hyperinsulinaemia, a feature of type 2 diabetes mellitus (T2DM), contributes to a cycle of events that may drive further insulin resistance (IR). ^1^ IR also appears to promote cardiovascular ^2^ and neurodegenerative diseases ^3^ while extreme IR represents a distinct category of T2DM ^4^. Lifestyle-based treatments are effective at reducing the risk of developing T2DM. ^5–8^ Such programs typically include prescription of physical activity and modify IR with high intra-individual variability. ^9–14^ Similar variability in IR responses to drug treatment also occur. ^15,16^ While monitoring the efficacy of any treatment on blood glucose is straightforward ^17^, the same cannot be said for monitoring insulin action. ^18,19^ Furthermore, the varying characteristics of commonly used insulin assays complicate comparisons across studies ^20,21^ while routine measurement of IR in clinical practice are considered too complex and costly. Established clinical definitions exist for T2DM and impaired glucose tolerance (IGT), but the definition of IR is less well developed. This is problematic because IR is distinct from IGT and may precede T2DM by decades. There is therefore a requirement to develop suitable technologies for diagnosing and tracking IR. ^22,23^ These IR biomarkers should offer practical (e.g. cost, simplicity) advantages over laboratory insulin assays. If such biomarkers can be shown to accurately predict longitudinal changes in IR, then efforts to configure low-cost standardised assays from minimally invasive sampling could be beneficial.

Interest in identification of robust and cost-effective IR diagnostics has led to the development of several clinical ^24^ and molecular prototypes including assays that measure branched-chain amino acids (BCAA)^25–28^, plasma lipoproteins^29–31^ and multi-protein signatures^32^ all of which correlate with IR status in cross-sectional or epidemiological cohorts. Useful biomarkers of IR should also reliably identify when a treatment for IR is ineffective, to allow the patient to be offered an alternative treatment. ^14–16,33,34^ There have been attempts to establish if plasma BCAA abundance tracks treatment responses yielding conflicting conclusions, partly because the studies were too small to reliably explore such relationships. ^35,36^ Determinants of insulin action combine multiple factors, including acute signalling mechanisms in the hours post-treatment (e.g. exercise) as well as more stable long-term adaptive changes ^37^ – such as increased tissue vascularisation. ^38^ Immediately post-exercise insulin action and glucose tolerance can be impaired, while insulin responses to an oral glucose tolerance test (OGTT) appear lowest 72hr following the previous exercise training session.^39^ Such temporal influences have not been considered when evaluating the available IR biomarkers and here we measured IR status within 24hr and 48-72hr of the final exercise training session. In the present study, we evaluated, in hundreds of individuals at-risk for T2DM, the performance of established BCAA and lipid biomarkers, combined with simple to measure clinical phenotypes to estimate fasting IR and predict changes in insulin following supervised lifestyle interventions.^9,12,40,41^ We established that choice of insulin assay is critical for capturing interactions with exercise status and, using Bayesian projective prediction, that robust plasma biomarkers of IR status in cross-sectional and prospective studies are unable to predict treatment responses following exercise-training based treatments.

## Materials and Methods

### Clinical Cohorts

A useful molecular biomarker should be able to track insulin resistance status and changes with a clinical intervention, regardless of the nature of the intervention. In the present modelling we contrast diverse life-style interventions in multiple studies, with comparable metabolic health and sedentary behaviours and comparable treatment effects. The META-PREDICT study was approved by local ethics committees at all trial centres (the University of Nottingham Medical School Ethics Committee: D8122011 BMS; the Regional Ethical Review Board Stockholm: 2012/753-31/2; the ethics committee of the municipality of Copenhagen and Frederiksberg in Denmark: H-3-2012-024; Comite etico de Investigacion Humana de la ULPGC: CEIH-2012-02; and the Loughborough University Ethics Approvals Human Participants Sub-Committee: 12/EM/0223). All complied with the 2008 Declaration of Helsinki. Both STRRIDE I and AT/RT study protocols were approved by the institutional review boards at Duke University and East Carolina University. The STRRIDE-PD study protocol was approved by the institutional review board at Duke University. All participants provided both verbal and signed written informed consent.

META-PREDICT (MP) cohort: This consisted of 189 active participants, recruited as previously described from 5 geographical regions across Europe.^12^ All clinical methods relied on cross-site standard operating procedures (SOPs), while insulin and metabolomic measures were performed in a single central laboratory. All participants were classified as sedentary (<600 metabolic equivalents (METs) min·wk^-1^) using a modified International Physical Activity Questionnaire,^42^ and had a fasting blood glucose level consistent with World Health Organisation (WHO) criteria for impaired fasting glucose (6.1 - 6.9 mmol·l^-1^), and/or a BMI >27 kg·m^-2^. Participants were excluded if they had evidence of active cardiovascular, cerebrovascular, respiratory, gastrointestinal or renal disease or had a history of malignancy, coagulation dysfunction, musculoskeletal or neurological disorders, recent steroid or hormone replacement therapy, or any condition requiring long-term drug prescriptions. Exercise training was supervised and consisted of three high intensity cycling sessions (5-by-1 minute at 125% VO_2_ max) per week for 6 weeks.^12^ Prior to the baseline assessments, participants were instructed to refrain from exercise for three days. Supine blood pressure (Omron M2, Omron Healthcare, Kyoto, Japan) and resting heart rate (RHR) were determined as the average of three consecutive measurements.

STRRIDE AT/RT (S2) cohort (NCT00275145): This study examined the independent and combined effects of aerobic and resistance exercise on cardiometabolic health in sedentary, overweight or obese adults with mild to moderate dyslipidaemia (LDL 3.37-4.92mmol/l or HDL ≤1.03mmol/l in males and ≤1.16mmol/l in females). Participants were randomized to one of three groups for eight months: 1) aerobic training only: 14 KKW at 65-80% VO_2peak_; 2) resistance training only: 3 days/week, 8 exercises, 3 sets/exercise, 8-12 repetitions/set; 3) full combination of the aerobic and resistance training programs. ^41^

STRRIDE PD (S-PD) cohort (NCT00962962): This study enrolled older sedentary, overweight or obese adults without or without impaired fasting glucose into one of four intervention groups for six months: 1) low amount/moderate intensity (10 KKW at 40-55% VO_2reserve_): 2) high amount/moderate intensity (16 KKW at 40-55% V O_2researve_); 3) high amount/vigorous intensity (16 KKW at 65-80% VO_2reserve_); 4) clinical lifestyle prescription, which included low amount/moderate intensity exercise of 10 KKW at 40-55% VO_2reserve_ plus diet to achieve 7% body weight loss (comparable to the Diabetes Prevention Program (DPP). ^40^

### Plasma sample glucose and insulin analysis

Blood glucose was analysed using a YSI 2300 STAT Plus glucose analyser (Yellow Springs Inc. Ohio, USA) in MP and S2, and with a Beckman-Coulter DxC600 clinical analyzer (Brea, CA, USA) in S-PD. The samples from all cohorts were analysed using high sensitivity insulin ELISA (Dako A/S, Sweden). Two levels of QC solutions were run for insulin. Coefficients of variation (CV) were acceptable; 4.68-8.03 % on both levels. In MP we also used an automated analyser (Immulite 2000) for analyses of insulin during an oral glucose tolerance test. To assure comparability of the results, we independently checked the assay performance for cross detection of insulin and C-peptide. The WHO standard for insulin and C-peptide were obtained from NIBSC (The National Institute for Biological Standards and Control). A dilution series for both analytes was prepared and run on Dako ELISA and Immulite 2000. The diluted WHO insulin standards were subjected to the same procedure as the samples.

### Plasma sample metabolomic analysis

Metabolomic analyses for MP has not been previously published (other than fasting glucose and insulin). Samples were randomized for sample preparation and analyses resulting in an equal distribution of all centres and possible responders and non-responders. We used hierarchical clustering of metabolomic variables to evaluate if there was an obvious systematic centre specific bias (**Figure S3**). Samples were taken after an overnight fast as arterialized venous blood and run in duplicate. Results within ±20% CV were accepted and if this limit was not met, the sample was rerun. Quality controls (QC) were run for all analyses and with each batch of sample preparation to ensure stable performance of the analysis procedure. QC for all analyses included in house controls made from pooled EDTA plasma from healthy controls. The following methods were used for each metabolite. Sample analysis used a Konelab 20XTi photospectrometer (Thermo Fisher Scientific, Thermo Electron Oy, Vantaa Finland) for high density lipoprotein HDL cholesterol, LDL cholesterol and triacylglycerides (TAG). Amino acids were analyzed using HPLC 2695 (Waters, Watford, UK) with online derivatization and a fluorescence detector 474 (Waters, Watford, UK) using a method described previously.^43,44^ Fatty acids were analyzed using ultra-high performance liquid chromatography (Ultimate 3000 UHPLC, Thermo Fisher Scientific, Germering, Germany) coupled to a TSQ Vantage with electrospray ionization (ESI) and triple quadrupole mass spectrometer (MS/MS) (Thermo Fisher Scientific, San Jose, CS, USA). For the STRRIDE cohorts, samples were analysed for plasma glucose concentration at the research site (Duke University, USA) using a Beckman–Coulter DxC600 clinical analyser (Brea, CA, United States). Plasma samples were analysed on 400 MHz nuclear magnetic resonance (NRM) profilers at LipoScience, now LabCorp (Morrisville, NC, United States), as previously described ^45^. The lipoprotein parameters and the BCAA were identified by retrospectively analysing digitally stored spectra using the newly developed NMR-based lipoprotein LP4 algorithm, which correlate with mass spectrometry methods.^46^

As distinct methodologies were utilised for the metabolomic analyses between MP and STRRIDE cohorts (e.g. mass spectroscopy versus NMR profiling) we explored the compatibility of the metabolomic data common to our diagnostic models. For S2 and S-PD, both mass spectroscopy and NMR methodologies were utilised for lipid related metabolomics and Bland-Altman analysis was performed to assess measurement agreement between HDL and TAG (**Figure S4-7**). There was high correlation between methods for HDL (S2 r^2^ 0.96, p<0.001; S-PD r^2^ 0.89, p<0.001) (**Figure S4A**, **Figure S5A**) and TAG (S2 r^2^ 0.99, p<0.001; S-PD r^2^ 0.95, p<0.001) (**Figure S6A, Figure S7A**). Bland-Altman analysis revealed a mean HDL measurement error of -0.066 mmol/L in S2 (**Figure S4B**) and -0.064 mmol/L in S-PD (**Figure S5B**) and mean TAG measurement error of +0.02mmol/L in S2 (**Figure S6B**) and +0.04mmol/L in S-PD (**Figure S7B**) with 95% of observations within 1.96 standard deviation of mean. Work by Wolak-Dinsmore *et al* demonstrated that NMR overestimates VAL and LEU by >25% (mean value) and underestimates ILE by 10-15% (mean value). While MS was used in MP, S2 and S-PD relied on NMR for BCAA quantification. ^46^ Based on these known systematic differences, modelling was performed independently in each cohort, using cross-validation methods.

### Statistical Modelling

Raw data from the clinical chemistry analyses was analysed with GraphPad Prism 5 (Software MacKiev, 2007, version 5.01) and more advanced analysis was accomplished using STATISTICA 10 (StatSoft Inc., 2011, version 10.0.228.2). All subsequent analysis was performed in R versions 4.1.1 and 4.3.2. We utilised conventional and Bayesian strategies to model data. Potentially predictive variables were selected based on known associations with IR and T2DM. Linear regression modelling was applied to investigate associations between dependent and independent variables. Briefly, after reducing each dataset to complete cases common variables were identified across all three cohorts. The criterion for considering a clinically useful variable was pragmatic – it had to be cheap and reliably measured in a primary care setting e.g. maximal aerobic capacity relates to IR status in some but not all cohorts, but it was not considered because of the time and cost required to measure it accurately. From the variables selected, some were dropped because of high collinearity (e.g. systolic blood pressure was retained in favour of diastolic blood pressure and mean arterial pressure in the Bayesian analysis). We removed participants with obesity class III (BMI >40kg/m^2^) for the Bayesian analysis and all continuous variables were normalised. The latter steps were carried out using the tidymodels in R (10.32614/CRAN.package.tidymodels) ^47^ and applied consistently across all datasets. Collinearity between variables was checked for each dataset separately **(Figure S8)**. Apart from BCAA in the MP dataset, there were no pairwise correlations with an absolute value >0.5. Linear models were built with student-t priors on both the intercept and slope coefficient terms (df=3, mean=0 and scale=4). These were mildly informative on the scale of the normalised data. More details are provided in the supplementary material. We built Bayesian multivariable linear models using the brms package to predict change in circulating insulin using relevant clinical variables, circulating BCAA levels and selective lipids.^48^ These full models were considered reference models. We then used projective prediction to identify a subset of variables with predictive performance as close as possible to that of the reference model.^49^ Briefly, projective prediction first generates a solution path - the variable ranking - for each sub model examined. Then a leave-one-out cross-validation process determines the predictive performance of each sub model along the predictor ranking. Data and the analysis script can be found at 10.5281/zenodo.13850947. ^50^

## Results

Demographic, blood and metabolomic data for the three independent cohorts – MP, S2 and S-PD – are presented in **Table 1**.

**Table 1.**
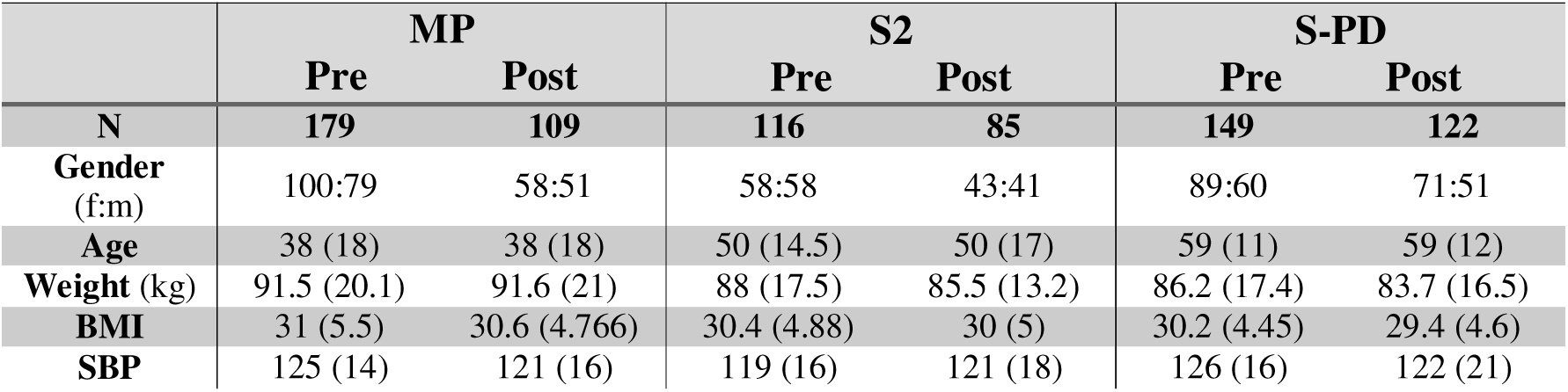
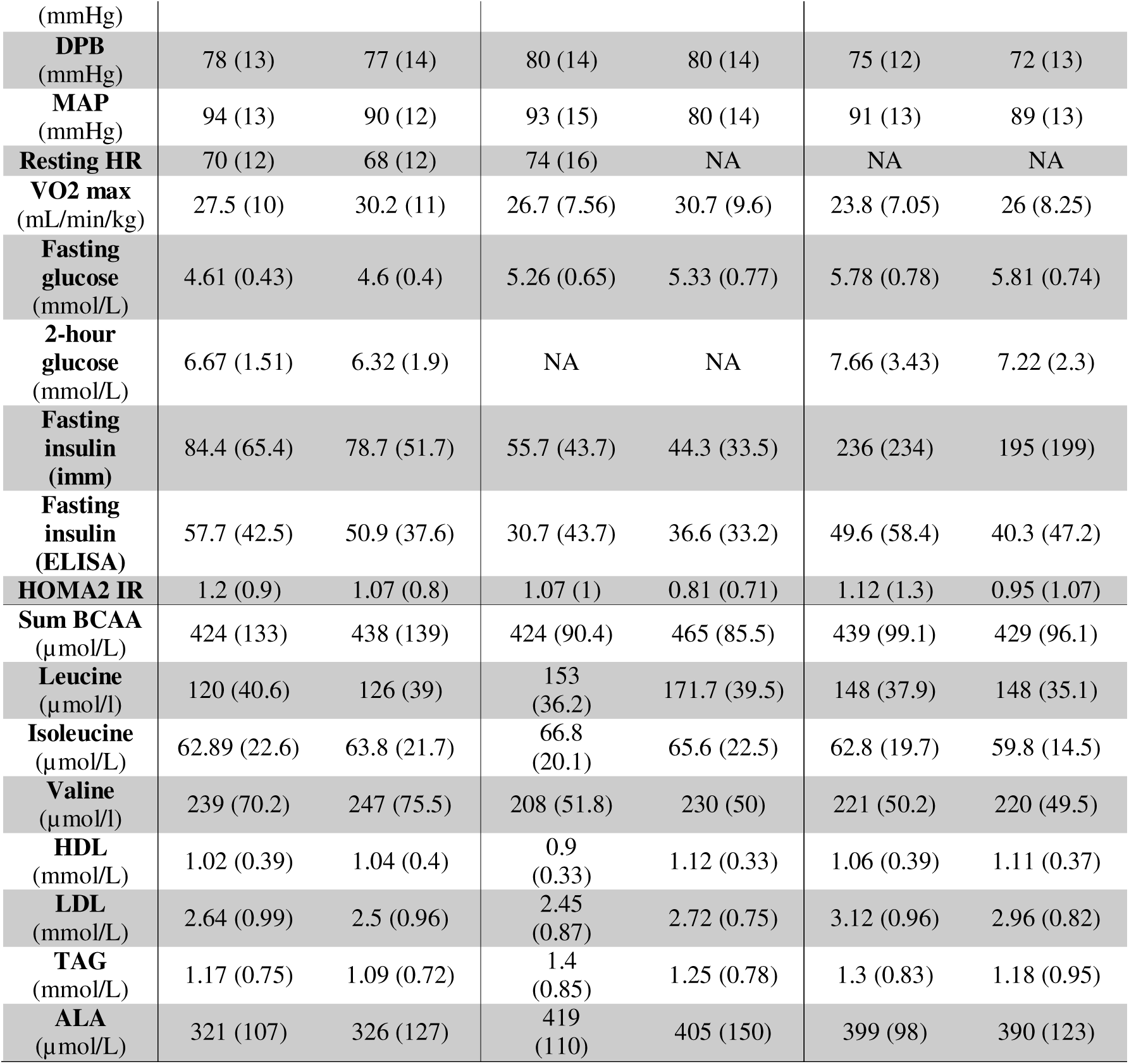
Demographics. Median and IQR. All available samples were utilised for baseline modelling. In each analysis, only subjects with complete data were used in the pre/post modelling, resulting in ∼ 20% fewer subjects than recruited or completing the intervention.

### Comparison of insulin assay and metabolomics assay performance

A long-standing challenge in studying the physiology of insulin is that commercial assays show distinct specificity and sensitivity profiles, with no agreement to move to a single standard assay.^20,21^ Certain insulin assays, utilised in exercise intervention studies (e.g. HERITAGE)^51^ or included in genome-wide association modelling,^52^ do not show molecular associations consistent with modern high sensitivity assays.^13^ In the present study we produced a large-scale comparison of two typical insulin assays (Immulite 2000) and a high sensitivity ELISA kit (Dako, Stockholm, Sweden), run concurrently in the same lab, on samples obtained before and after exercise-training that reduced IR.^53^ Both Immulite 2000 and Dako ELISA showed a strong correlation between expected and measured insulin; R^2^=0.999 and R^2^=0.992, respectively (International WHO standard for insulin, The National Institute for Biological Standards and Control, **Figure S1**). No cross reaction between insulin and c-peptide standards was detected (data not shown). When plotting both insulin values from the same fasting samples, there was moderate agreement between the two assays (R^2^=0.65, **Figure S2)**. Critically, using insulin measurements from the high-sensitivity ELISA, we were able to detect a significant reduction in IR following 6 weeks exercise, while the Immulite 2000 immune-assay was unable to detect a change (**Figure 1**). All the subsequent analyses in this article relied on insulin values obtained from the Dako high-sensitivity ELISA (produced in a single laboratory).

**Figure 1.**
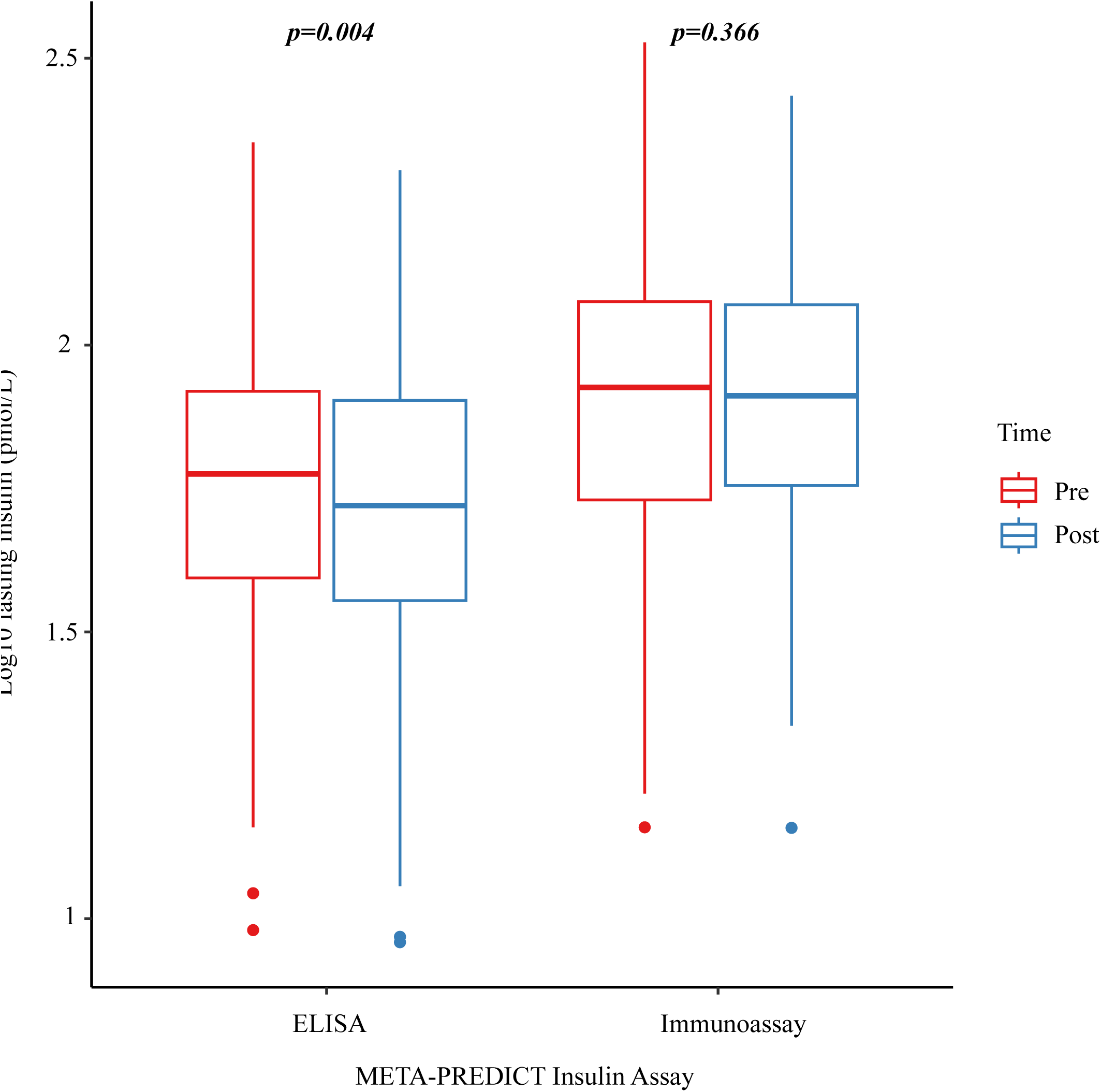
Insulin assay statistical performance for identifying a statistically significant difference in pre vs post exercise training samples from all included exercise protocols. Fasting insulin (pmol/L) was measured using a high sensitivity ELISA (right) and the Immulite 2000 automated analyser (left). P-values are calculated from paired t-tests on log10 fasting insulin (pmol/L).

### Biomarker association with fasting insulin and HOMA2-IR status

Potential predictor variables were selected based on the literature summarised above and availability in all three cohorts. Univariable linear regression analysis was performed to explore individual relationships between fasting insulin and the selected clinical and metabolomic biomarkers (**Figure 2**; full results in **Table S1**). In MP, all pre-selected variables except age (p=0.09) reached statistical significance for linear association with fasting insulin (age is a reliable covariate but the age-range in MP was limited). In S2 log_10_ BMI (p<0.001), fasting glucose (p<0.001), HDL (p=0.007), TAG (p<0.001) and amino acids (isoleucine (ILE) p=0.0005, leucine (LEU) p=0.006, valine (VAL) p<0.001, alanine (ALA) p=0.0027) were significant for a linear association with fasting insulin. Univariate associations were very similar for S-PD except ALA (p=0.068). For subsequent integrated models and treatment responses predictions, variables which showed a statistically significant association in one or more of the cohorts were considered further (given each was already supported with previously published evidence for having a cross-sectional association with IR status).

**Figure 2.**
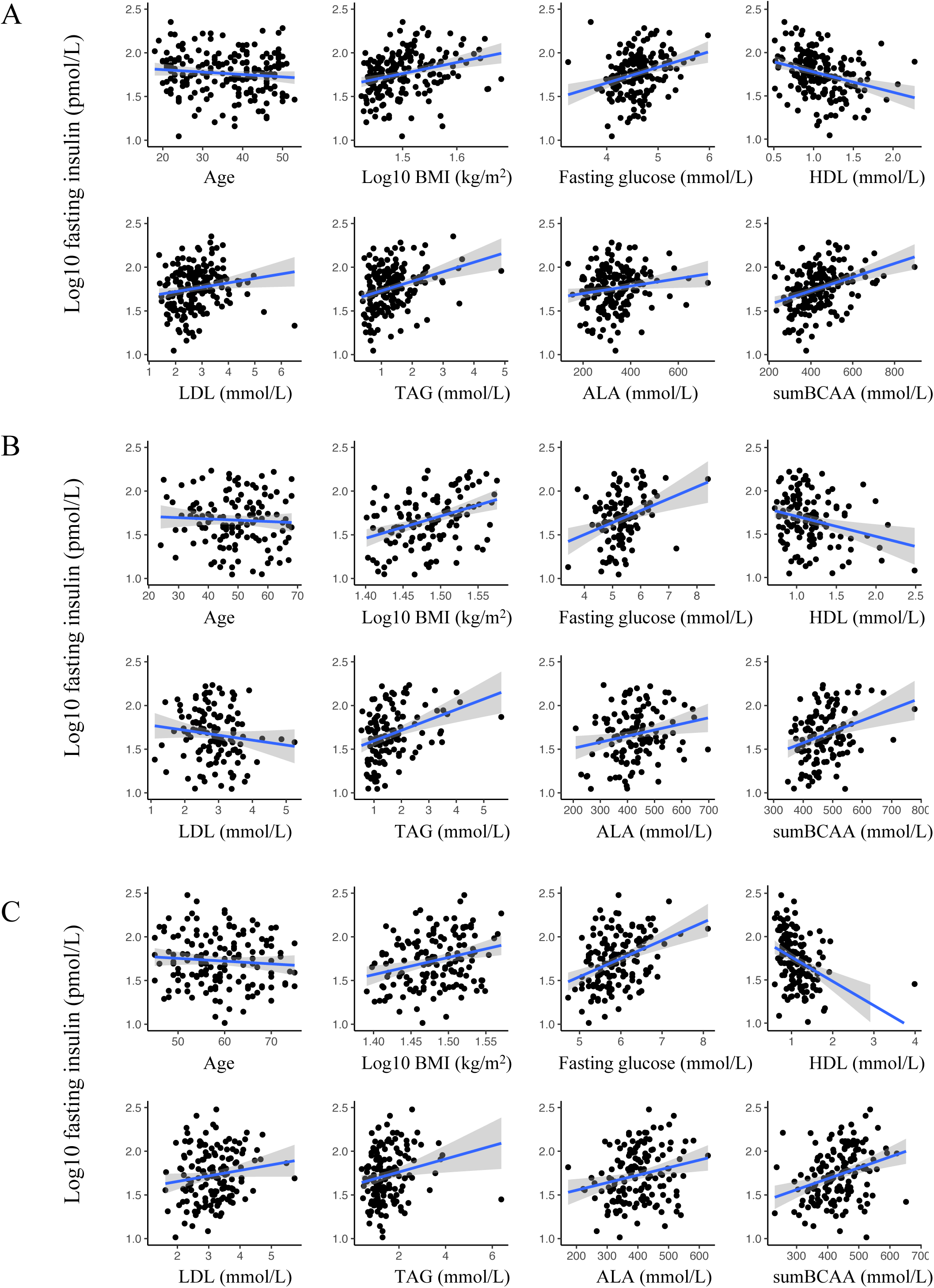
Scatter plots with OLS line of best fit (95% CI shaded) between log10 fasting insulin and clinical and metabolic variables. A) MP, B) S2 and C) S-PD

As each study used distinct metabolomic methods (see Methods) we performed multivariable linear regression and logistic regression with K-fold cross validation (k=10) separately in each cohort. **Table S2** demonstrates the same models discussed below, to predict fasting insulin using K-fold cross validation multiple linear regression, with the combined model demonstrating the strongest association with directly measured insulin (r^2^ 0.33 – 0.39). Fasting glucose was excluded when using HOMA2-IR as the dependant variable due to its inclusion in the HOMA2-IR model. To evaluate classification performance, cohorts were divided into insulin sensitive (HOMA2-IR <1.3) and insulin resistant (HOMA2-IR ≥1.3) based on values obtained in a large population study (n=95,540), where a HOMA2-IR ≥1.3 was associated with a hazard ratio of 3.2 (95% CI 1.9-5.3) for the development of T2DM over a median of 4.7y. ^54^ The non-diabetic subjects in this study (n=93,710) are Caucasians living in Denmark. They were of similar age and metabolic health to the STRRIDE cohorts described in this study (84.8% of S2 and 78.2% of S-PD were Caucasian). The MP cohort are of the similar ethnicity and metabolic health to the population study but were younger (median age 38 vs 57) with a higher BMI (median BMI 31 vs 26). Four models were assessed in each cohort (**Figure 3**); a baseline model (age, BMI and gender), a BCAA model (age, BMI, gender and sum of BCAA), a lipid model (age, BMI, gender, HDL, LDL and TAG) and finally, a combined model (age, BMI, gender, BCAA, HDL, LDL and TAG). This allowed us to evaluate if lipids (HDL, LDL and TAG) and BCAA (ILE, LEU and VAL) add value to risk estimation from simple clinical variables. The baseline (‘clinical’) model had no discriminatory performance in S-PD, so the addition of metabolomic variables improved the model substantially. The statistical advantage of including metabolites in the model applied to S-PD may reflect the more homogeneous nature of that cohort, for the utilised clinical parameters. Otherwise, inclusion of BCCA and/or lipids offered no meaningful value, with poor sensitivity (**Table 2)**.

**Figure 3.**
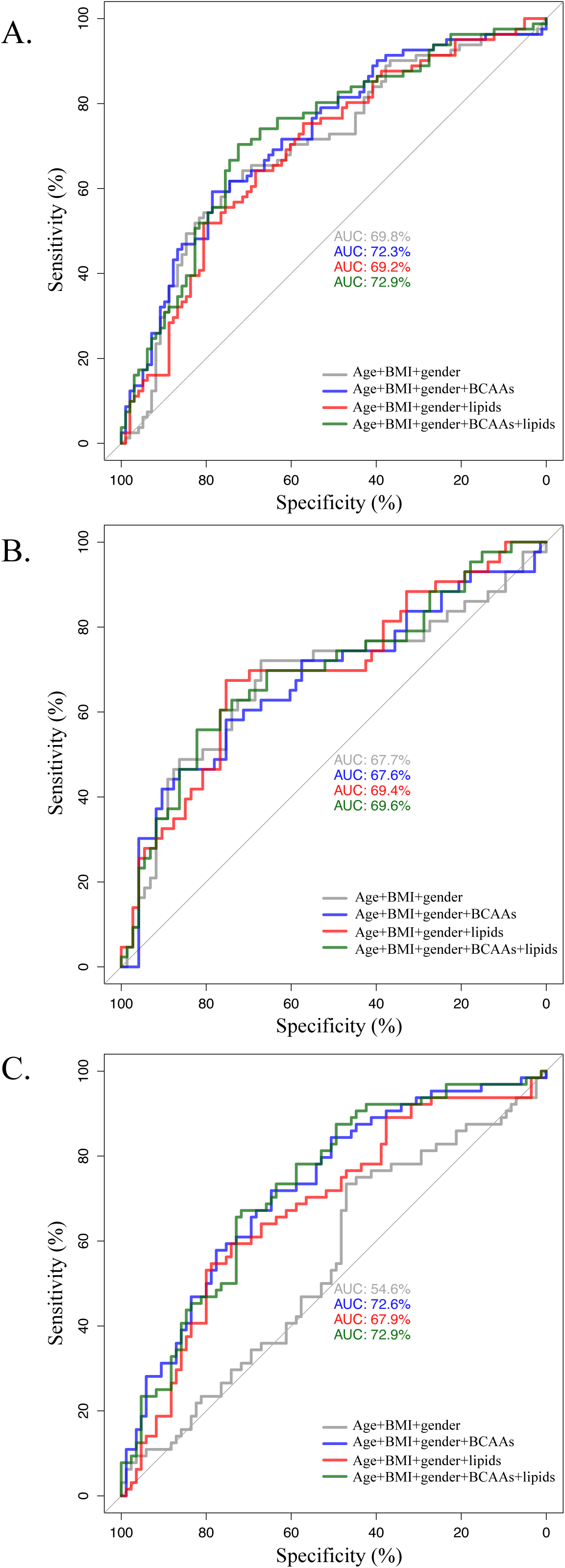
Utility of baseline c linical and metabolomic variables to classify HOMA2-IR status at 1.3 unit threshold reflecting the analysis of Marott et al.^55^ K-fold cross validation logistic regression ROC curves for A. MP, B. S2 and C. S-PD.

**Table 2.** K-fold cross validation logistic regression model statistics for combined model predicting insulin resistance (HOMA2 IR ≥1.3) using age, BMI, gender, sum of branched chain amino acids, HDL, LDL and triglycerides.

|  | **MP** | **S2** | **S-PD** |
| --- | --- | --- | --- |
| **Accuracy (95% CI)** | 0.676 (0.602-0.744) | 0.707 (0.615-0.788) | 0.644 (0.562-0.721) |
| **Kappa** | 0.339 | 0.347 | 0.27 |
| **Sensitivity** | 0.58 | 0.512 | 0.563 |
| **Specificity** | 0.755 | 0.822 | 0.706 |
| **PPV** | 0.662 | 0.629 | 0.59 |
| **NPV** | 0.685 | 0.741 | 0.682 |
| **Prevalence** | 0.453 | 0.371 | 0.43 |
| **Detection rate** | 0.263 | 0.19 | 0.242 |

### Biomarker based prediction of IR status in response to lifestyle intervention

The results above established that each potential biomarker associates with fasting insulin in some or all trials (**Table S1**) but that multivariable models do not identify *IR status* (based on a HOMA2-IR threshold) in any cohort **(Figure 3**, **Table 2)**. The utility of the same biomarkers for *predicting* exercise treatment related insulin responses was now examined. For this we included changes in fasting glucose (as it is easy and inexpensive to measure). **Figure 4** displays the univariable linear relationships of changes in clinical and metabolomic biomarkers with changes in fasting insulin as measured with the high sensitivity ELISA. **Figure S8** shows the same data but displaying correlation coefficients (including individual amino acids). To further examine the *predictive* utility of individual and combinations of biomarkers we built Bayesian linear models independently for each cohort modelling change in circulating insulin after the exercise interventions used. These models included age and changes over the intervention in individual BCAA, fasting glucose, BMI, lipids, systolic blood pressure and alanine. The full models (“reference” models) were considered as the optimum solution, given the present data, to the prediction task.^55^ The posterior predictive distributions showed the models performed well (**Fig 5A-C**). Using projective prediction, we found that the biomarkers did not improve predictive performance over a null model i.e. the intercept term (**Figure 5D-F**).^49^ Thus, change in biomarkers (alone or additively combined) did not predict change in insulin following an exercise intervention.

**Figure 4.**
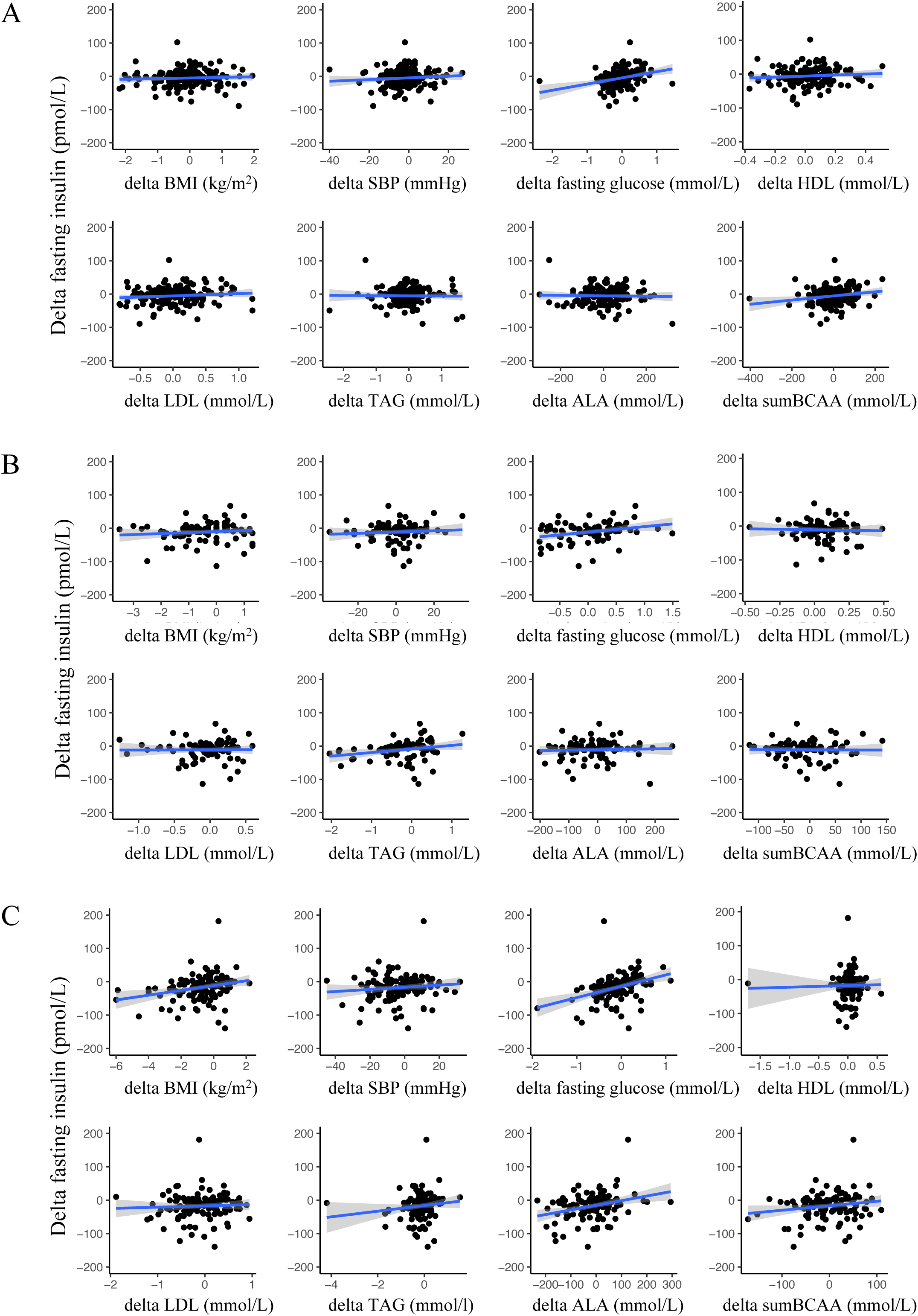
Scatter plots with OLS line of best fit (95% CI shaded) between delta fasting insulin and clinical and metabolic variables. A) MP, B) S2 and C) S-PD

**Figure 5.**
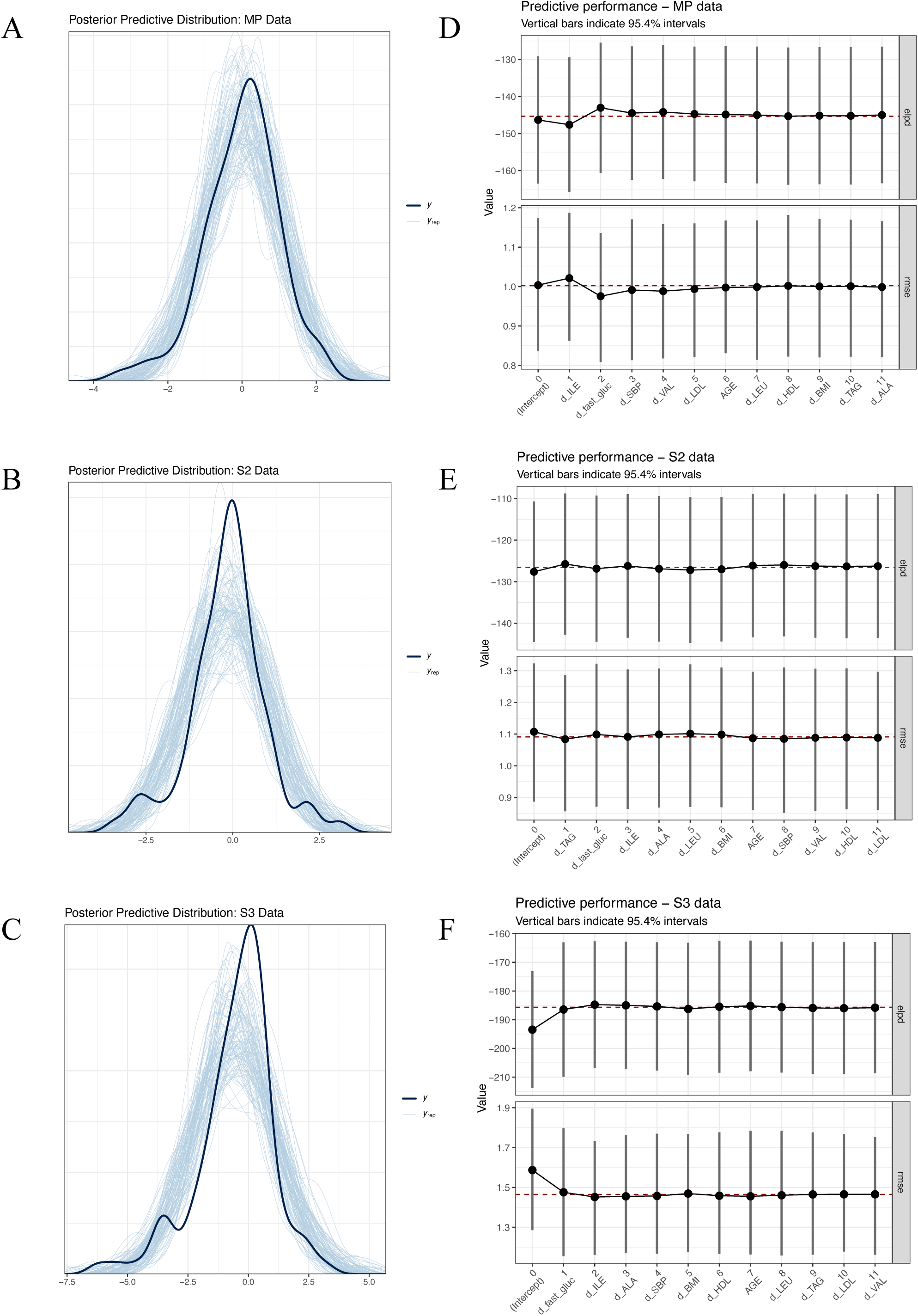
Left panel; Posterior predictive distributions for A. MP, B. S2 and C. S-PD. Right panel; Expected log predictive density (Elpd, upper trace) and root mean square error (RMSE, lower trace) for D. MP, E. S2 and F. S-PD. Elpd and RMSE are shown with 95% nominal coverage intervals.

## Discussion

Development of robust and cost-effective diagnostics should contribute to combating the T2DM epidemic and facilitate alignment of clinical practice with the recommendations of the International Diabetes Federation, namely earlier detection and prevention of pre-diabetes.^56^ Insulin is not routinely measured in clinical practice and thus crucial information, regarding the temporal characteristics of IR over disease development, is lacking. Previous work has identified several potential biomarkers for IR in cross-sectional and prospective settings. ^25–27,29–31,57^ The present study extends this body of work to evaluate these IR biomarkers in the context of changes in circulating insulin in response to life-style intervention, including exercise training.

### Importance of the insulin assay

To our knowledge we are the first to study the importance of insulin assay performance on tracking insulin changes in response to exercise interventions. The MP cohort represents typical younger-middle aged individuals with multiple risk factors for T2DM.^12^ We found that the Immulite 2000 immunoassay was unable to detect changes in fasting insulin status following supervised exercise training.12 In contrast, concurrent measurements using a high sensitivity ELISA assay detected a significant reduction in fasting insulin. Neither assay demonstrated any cross-reactivity with c-peptide and a strong correlation with the WHO standard. The American Diabetes Association has attempted to promote a consensus on standardizing the measurement of insulin.^20,21^ Our analysis indicates that when environmental influences are to be considered a high-sensitivity ELISA insulin assay must be utilised. We have previously observed that some older insulin assays, with unclear specificity^58^ were unsuitable for genomic association analysis.^13^ Indeed, it is plausible that the use of insulin assays insensitive to variations within the ‘normal’ range, or with some specificity for C-peptide, limited genome-wide association modelling of fasting insulin.^59^ While suboptimal insulin assays will not be voluntarily retired by manufacturers, it would seem sensible that the research community adopt the use of a limited number of high-sensitivity assays for studies of fasting insulin in most situations^20,21^ especially when subtle gene-environment interactions are being modelled.

### Metabolomic components of the IR biomarker models

There is a well-established positive correlation between individual blood BCAA and IR. ^25–27^ As such, pathways that control BCAA catabolism have gained interest as possible targets for reversing IR.^28,60,61^ Furthermore, BCAA have been combined with clinical parameters to increase the performance of models that predict both IR and risk of T2DM.^29,31,57^ We previously identified, in four independent cohorts, a transcriptional signature including genes controlling BCAA catabolism pathways, that related to IR after adjustment for age, VO2max and BMI (n=564). We also found that in response to four independent lifestyle interventions (n=196), expression of 16 genes in human skeletal muscle changed in proportion to improvements in insulin sensitivity, with 25% of these related to BCAA metabolism.^13^ By mass, skeletal muscle has the largest capacity for BCAA catabolism in humans supporting the plausibility of these observations.^62^

We found plasma BCAA were unable to predict changes in IR status in the present study. Insulin mediated BCAA clearance, however, may rely on other organs including the liver meaning that blood levels may not accurately reflect metabolism of BCCA.^63^ In a recent study, Lee et al reported that baseline plasma BCAA levels related to change in glucose infusion rate during a hyperinsulinaemic−euglycaemic clamp as a measure of insulin sensitivity and liver fat content.^36^ This association occurred despite no change in total plasma BCAA levels and a reported increase in muscle BCAA catabolism pathway gene expression. We also found no change in BCAA levels with any exercise training protocol (**Table 1**), which might be understandable as only ∼40% of subjects demonstrate a numerical improvement in HOMA2-IR while some demonstrate an increase in IR.^12,13^ Our observations are consistent with earlier distinct modelling approaches applied to the STRRIDE cohorts where the combination of BCAA and lipid-based IR biomarkers did not reliably detect changes in insulin sensitivity produced by a combination of resistance and aerobic training.^17^ Various other combinations of BCAA and protein metabolites have been considered in IR models and while some variation is observed,^64^ these are likely random associations found in small cohorts. Using energy restriction to modify IR, we recently reported that group mean differences in plasma BCAA abundance were unchanged, despite group mean improvements in IR. ^64^ These observations contrast with a previous study, relying on a sample size 10 times smaller, where changes in plasma BCAA abundance *did* track with improved IR status.^35^

In the present analysis we considered several strategies, including several distinct clinical and metabolomic variables, to model both fasting insulin and HOMA2-IR. A practical approach would combine informative but easy to measure (cost effective) clinical variables with metabolomic markers (that could be measured in the future using cost-effective devices). Bayesian modelling and projective prediction ^65,66^ was used to establish a sparse model that retained the ability to provide useful predictions of circulating insulin changes. Projective prediction uses the predictions from the reference (full) model to identify a subset of variables with predictive ability as close as possible to that of the full model. This was unable to identify a smaller set of variables (or indeed any variables) useful for predicting change in circulating insulin after lifestyle intervention using this strategy. Conventional regression modelling, against changes in HOMA2-IR also failed to identify a useful model.

### Clinical evaluation of IR biomarker models

Diagnosing IR is challenging not least because the definitions of being “IR” remains imprecise. The gold standard method is commonly claimed to be the ‘clamp’ technique or intravenous glucose tolerance test (IGTT).^19,67^ However, this requires substantial resources and therefore the IGTT is rarely used for diagnosis. The ‘clamp’ by its nature is non-physiological (e.g. individuals can be exposed to non-physiological conditions). Alternative indices of IR and pancreatic beta cell function exist and include the homeostatic model assessment (HOMA) and HOMA2 ^68,69^, Matsuda index ^70^ and the disposition index (DI).^71^ These indices are also not used widely, again largely because insulin is not routinely measured, a fact that in turn prevents establishment of reliable thresholds for diagnosis of IR. Although several epidemiological studies have used HOMA and HOMA2, there is limited modelling to define a clinically useful cut-off value for IR. Several of these studies used a percentile cut off and then applied ROC analysis to determine the threshold for a diagnosis of IR. This was then related to relative risk for relevant clinical outcomes e.g. progression to T2DM or cardiovascular disease. As a result, the HOMA-IR threshold reported to define ‘at risk’ varies from 1.8 to 3.9 depending on cohort characteristics and methods used.^18^ One very large study estimated that a HOMA2-IR in excess of 1.3 represented a hazard ratio of >3.2 for developing T2DM over an average of ∼5 years in 95,450 subjects.^54^ This is the largest study linking a particular HOMA2-IR value to incipient diabetes and we used this HOMA2-IR cut-off in the present study. Interestingly, in exploratory analysis we conducted, 83% of those with the most severe IR (HOMA2-IR ≥ 2.4) were not classified as metabolically compromised based on fasting glucose or 2-hour-glucose OGTT data. This illustrates that the current reliance on glucose-centric risk assessment limits our ability to identify (and hence intervene) in those people with IR. Nevertheless, all surrogate variables investigated in the present study had only a modest ability to diagnose IR in a cross-sectional setting, and no ability to track changes in IR with exercise or diet. Thus, new large-scale trials, where IR is treated by various means, and insulin and biomarker measurements are made, are urgently required.

In summary, we evaluated multiple novel predictive models for IR, including models incorporating cholesterol species and BCAA. These models failed to predict change in IR status following supervised lifestyle modification using multiple interventions. A main limitation of our study is the limited availability of common metabolomic variables across cohorts. Nevertheless, we are not aware of any blood based metabolic disease biomarkers that sensitively track improvements in insulin sensitivity across multiple independent cohorts.

## Data Availability Statement

A majority of the datasets generated during and/or analyzed during the current investigation are not publicly available but are available from the corresponding author on reasonable request.

## Contributors

RJB, JAT, OR and IJG conceptualized aims and designed the analysis strategy. All authors contributed to data collection and/or pre-processing. RJB and IJG performed data analysis and JAT contributed to interpretation. JAT and RJB wrote the first draft and JAT, RJB and IJG produced a full manuscript. All authors contributed to revision of manuscript and have read and approved the final manuscript.

## Figure Legends

**Figure S1.** Performance of the insulin assays. A) Immulite using titration international insulin standard, B) ELISA using titration international insulin standard, and C) Immulite vs ELISA standard curve values using international insulin standard once converted to uIU/ml.

**Figure S2.** Correlation of participant fasting insulin measured using Dako ELISA and Immulite 2000 assays.

**Figure S3.** Hierarchical clustering of subjects by their metabolomic profile demonstrating that there is no centre specific bias. Copenhagen (CPH), Las Palmas (LPA), Loughborough (LU), Stockholm (Stock).

**Figure S4.** S2 HDL assay performance. A) Comparison of mass spectroscopy and nuclear magnetic resonance (NMR) measurement of HDL using linear regression. B) Bland-Altman plot to estimate of bias with 95% confidence interval shown in blue, with upper (green) and lower (red) limits.

**Figure S5.** S-PD HDL assay performance. A) Comparison of mass spectroscopy and nuclear magnetic resonance (NMR) measurement of HDL using linear regression. B) Bland-Altman plot to estimate of bias with 95% confidence interval shown in blue, with upper (green) and lower (red) limits.

**Figure S6.** S2 TAG assay performance. A) Comparison of mass spectroscopy and nuclear magnetic resonance (NMR) measurement of TAG using linear regression. B) Bland-Altman plot to estimate of bias with 95% confidence interval shown in blue, with upper (green) and lower (red) limits.

**Figure S7.** S-PD TAG assay performance. A) Comparison of mass spectroscopy and nuclear magnetic resonance (NMR) measurement of TAG using linear regression. B) Bland-Altman plot to estimate of bias with 95% confidence interval shown in blue, with upper (green) and lower (red) limits.

**Figure S8.** Correlation plots showing strength of correlation between the variables used in the generation of linear models to predict change in circulating insulin across lifestyle interventions. A. MP, B. S2 and C. S-PD

## Tables

**Table S1.** Linear regression modelling using log10 fasting insulin as dependant variable. Note the sample sizes reflecting all subjects with data for each individual clinical parameter or metabolomic measure. In final integrated modelling only subjects with all completes values were included.

**Table S2.** K-fold cross validation multiple linear regression predicting baseline log_10_ fasting insulin from clinical and metabolomic variables

## References

1. Cen HH, Hussein B, Botezelli JD, et al. Human and mouse muscle transcriptomic analyses identify insulin receptor mRNA downregulation in hyperinsulinemia-associated insulin resistance. FASEB Journal. 2022;36(1).

2. Wamil M, Coleman RL, Adler AI, McMurray JJV, Holman RR. Increased Risk of Incident Heart Failure and Death Is Associated With Insulin Resistance in People With Newly Diagnosed Type 2 Diabetes: UKPDS 89. Diabetes Care. 2021;44(8):1877–1884.

3. Folch J, Olloquequi J, Ettcheto M, et al. The Involvement of Peripheral and Brain Insulin Resistance in Late Onset Alzheimer’s Dementia. Front Aging Neurosci. 2019;11:236.

4. Dennis JM, Shields BM, Henley WE, Jones AG, Hattersley AT. Disease progression and treatment response in data-driven subgroups of type 2 diabetes compared with models based on simple clinical features: an analysis using clinical trial data. Lancet Diabetes Endocrinol. 2019;7(6):442–451.

5. Knowler WC, Fowler SE, Hamman RF, et al. 10-year follow-up of diabetes incidence and weight loss in the Diabetes Prevention Program Outcomes Study. Lancet. 2009;374(9702):1677-1686.

6. Wing RR, Bolin P, Brancati FL, et al. Cardiovascular effects of intensive lifestyle intervention in type 2 diabetes. New England Journal of Medicine. 2013;369(2):145–154.

7. Uusitupa M, Peltonen M, Lindström J, et al. Ten-year mortality and cardiovascular morbidity in the Finnish Diabetes Prevention Study - Secondary analysis of the randomized trial. PLoS One. 2009;4(5):1–8.

8. Hivert MF, Christophi CA, Franks PW, et al. Lifestyle and metformin ameliorate insulin sensitivity independently of the genetic burden of established insulin resistance variants in diabetes prevention program participants. Diabetes. 2016;65(2):520–526.

9. Ross LM, Slentz CA, Kraus WE. Evaluating Individual Level Responses to Exercise for Health Outcomes in Overweight or Obese Adults. Front Physiol. 2019;10.

10. Huffman KM, Slentz CA, Bateman LA, et al. Exercise-induced changes in metabolic intermediates, hormones, and inflammatory markers associated with improvements in insulin sensitivity. Diabetes Care. 2011;34(1):174–176.

11. AbouAssi H, Slentz CA, Mikus CR, et al. The Effects of Aerobic, Resistance and Combination Training on Insulin Sensitivity and secretion in Overweight Adults from STRRIDE AT/RT: A Randomized Trial. J Appl Physiol (1985). 2015;118(12)1474-1482.

12. Phillips BE, Kelly BM, Lilja M, et al. A practical and time-efficient high-intensity interval training program modifies cardio-metabolic risk factors in adults with risk factors for type II diabetes. Front Endocrinol (Lausanne). 2017;8:229.

13. Timmons JA, Atherton PJ, Larsson O, et al. A coding and non-coding transcriptomic perspective on the genomics of human metabolic disease. Nucleic Acids Res. 2018;46(15):7772–7792.

14. Álvarez C, Ramírez-Campillo R, Ramírez-Vélez R, Izquierdo M. Effects and prevalence of nonresponders after 12 weeks of high-intensity interval or resistance training in women with insulin resistance: a randomized trial. J Appl Physiol. 2017;122:985–996.

15. Sears DD, Hsiao G, Hsiao A, et al. Mechanisms of human insulin resistance and thiazolidinedione-mediated insulin sensitization. Proc Natl Acad Sci U S A. 2009;106(44):18745–18750.

16. Brivba M, Ansone L, Silamikelis I, et al. Whole-blood transcriptome profiling reveals signatures of metformin and its therapeutic response. PLoS One. 2020;15(8):e0237400.

17. Ross LM, Slentz CA, Zidek AM, et al. Effects of Amount, Intensity, and Mode of Exercise Training on Insulin Resistance and Type 2 Diabetes Risk in the STRRIDE Randomized Trials. Front Physiol. 2021;12:1–10.

18. Tang Q, Li X, Song P, Xu L. Optimal cut-off values for the homeostasis model assessment of insulin resistance (HOMA-IR) and pre-diabetes screening: Developments in research and prospects for the future. Drug Discov Ther. 2015;9(6):380–385.

19. Ferrannini E, Mari A. How to measure insulin sensitivity. J Hypertens. 1998;16(7):895–906.

20. Manley SE, Stratton IM, Clark PM, Luzio SD. Comparison of II human insulin assays: Implications for clinical investigation and research. Clin Chem. 2007;53(5):922–932.

21. Miller WG, Thienpont LM, Van Uytfanghe K, et al. Toward standardization of insulin immunoassays. Clin Chem. 2009;55(5):1011–1018.

22. Thambisetty M, Metter EJ, Yang A, et al. Glucose intolerance, insulin resistance, and pathological features of Alzheimer disease in the Baltimore Longitudinal Study of Aging. JAMA Neurol. 2013;70(9):1167–1172.

23. Rodríguez-Mañas L, Angulo J, Carnicero JA, El Assar M, García-García FJ, Sinclair AJ. Dual effects of insulin resistance on mortality and function in non-diabetic older adults: findings from the Toledo Study of Healthy Aging. Geroscience. 2022;44(2):1095–1108.

24. Clausen JO, Borch-Johnsen K, Ibsen H, et al. Insulin sensitivity index, acute insulin response, and glucose effectiveness in a population-based sample of 380 young healthy Caucasians: Analysis of the impact of gender, body fat, physical fitness, and life-style factors. Journal of Clinical Investigation. 1996;98(5):1195–1209.

25. Zheng Y, Ceglarek U, Huang T, et al. Weight-loss diets and 2-y changes in circulating amino acids in 2 randomized intervention trials. American Journal of Clinical Nutrition. 2016;103(2):505–511.

26. Shah SH, Bain JR, Muehlbauer MJ, et al. Association of a peripheral blood metabolic profile with coronary artery disease and risk of subsequent cardiovascular events. Circ Cardiovasc Genet. 2010;3(2):207–214.

27. Bloomgarden Z. Diabetes and branched-chain amino acids: What is the link? J Diabetes. 2018;10(5):350–352.

28. Newgard CB, An J, Bain JR, et al. A branched-chain amino acid-related metabolic signature that differentiates obese and lean humans and contributes to insulin resistance. Cell Metab. 2009;9(4):311–326.

29. Floegel A, Stefan N, Yu Z, et al. Identification of serum metabolites associated with risk of type 2 diabetes using a targeted metabolomic approach. Diabetes. 2013;62(2):639–648.

30. Shalaurova I, Connelly MA, Garvey WT, Otvos JD. Lipoprotein insulin resistance index: a lipoprotein particle-derived measure of insulin resistance. Metab Syndr Relat Disord. 2014;12(8):422–429.

31. Flores-Guerrero JL, Gruppen EG, Connelly MA, et al. A newly developed diabetes risk index, based on lipoprotein subfractions and branched chain amino acids, is associated with incident type 2 diabetes mellitus in the prevend cohort. J Clin Med. 2020;9(9):1–17.

32. Zanetti D, Stell L, Gustafsson S, et al. Plasma proteomic signatures of a direct measure of insulin sensitivity in two population cohorts. Diabetologia. 2023;66(9):1643–1654.

33. Mutch DM, Temanni MR, Henegar C, et al. Adipose gene expression prior to weight loss can differentiate and weakly predict dietary responders. PLoS One. 2007;2(12).

34. Newton RL, Johnson WD, Larrivee S, et al. A Randomized Community-based Exercise Training Trial in African American Men: Aerobic Plus Resistance Training and Insulin Sensitivity in African American Men. Med Sci Sports Exerc. 2020;52(2):408–416.

35. Glynn EL, Piner LW, Huffman KM, et al. Impact of combined resistance and aerobic exercise training on branched-chain amino acid turnover, glycine metabolism and insulin sensitivity in overweight humans. Diabetologia. 2015;58(10):2324–2335.

36. Lee S, Gulseth HL, Langleite TM, et al. Branched-chain amino acid metabolism, insulin sensitivity and liver fat response to exercise training in sedentary dysglycaemic and normoglycaemic men. Diabetologia. 2021;64(2):410–423.

37. Dimenna FJ, Arad AD. The acute vs. chronic effect of exercise on insulin sensitivity: nothing lasts forever. Cardiovasc Endocrinol Metab. 2021;10(3):149–161.

38. Zheng C, Liu Z. Vascular function, insulin action, and exercise: an intricate interplay. Trends Endocrinol Metab. 2015;26(6):297–304.

39. King DS, Baldus PJ, Sharp RL, Kesl LD, Feltmeyer TL, Riddle MS. Time course for exercise-induced alterations in insulin action and glucose tolerance in middle-aged people. J Appl Physiol (1985). 1995;78(1):17-22.

40. Slentz CA, Bateman LA, Willis LH, et al. Effects of exercise training alone vs a combined exercise and nutritional lifestyle intervention on glucose homeostasis in prediabetic individuals: a randomised controlled trial. Diabetologia. 2016;59(10):2088–2098.

41. Slentz CA, Bateman LA, Willis LH, et al. Effects of aerobic vs. resistance training on visceral and liver fat stores, liver enzymes, and insulin resistance by HOMA in overweight adults from STRRIDE AT/RT. Am J Physiol Endocrinol Metab. 2011;301(5):1033–1039.

42. Hagströmer M, Oja P, Sjöström M. The International Physical Activity Questionnaire (IPAQ): a study of concurrent and construct validity. Public Health Nutr. 2006;9(6):755–762.

43. Godel H, Graser T, Földi P, Pfaender P, Fürst P. Measurement of free amino acids in human biological fluids by high-performance liquid chromatography. J Chromatogr A. 1984;297:49–61.

44. Vesali RF, Klaude M, Rooyackers O, Wernerman J. Amino acid metabolism in leg muscle after an endotoxin injection in healthy volunteers. Am J Physiol Endocrinol Metab. 2005;288:360–364.

45. Jeyarajah EJ, Cromwell WC, Otvos JD. Lipoprotein particle analysis by nuclear magnetic resonance spectroscopy. Clin Lab Med. 2006;26(4):847–870.

46. Wolak-Dinsmore J, Gruppen EG, Shalaurova I, et al. A novel NMR-based assay to measure circulating concentrations of branched-chain amino acids: Elevation in subjects with type 2 diabetes mellitus and association with carotid intima media thickness. Clin Biochem. 2018;54:92–99.

47. Kuhn M, Wickham H. Tidymodels: a collection of packages for modeling and machine learning using tidyverse principles. Published 2020. Accessed April 04, 2025. https://www.tidymodels.org

48. Bürkner PC. brms: An R package for Bayesian multilevel models using Stan. J Stat Softw. 2017;80.

49. Piironen J, Paasiniemi M, Vehtari A. Projective inference in high-dimensional problems: Prediction and feature selection. Electron J Stat. 2020;14(1):2155–2197.

50. Gallagher I, Timmons J, Brogan R. Biomarkers of insulin resistance and their performance as predictors of treatment response in adults with risk factors for type 2 diabetes. Zenodo. Accessed April 04, 2025 10.5281/zenodo.13819578

51. An P, Teran-Garcia M, Rice T, et al. Genome-wide linkage scans for prediabetes phenotypes in response to 20 weeks of endurance exercise training in non-diabetic whites and blacks: The HERITAGE Family Study. Diabetologia. 2005;48(6):1142–1149.

52. Bouchard C, Rankinen T, Timmons JA. Genomics and genetics in the biology of adaptation to exercise. Compr Physiol. 2011;1(3):1603–1648.

53. Phillips BE, Kelly BM, Lilja M, et al. A practical and time-efficient high-intensity interval training program modifies cardio-metabolic risk factors in adults with risk factors for type II diabetes. Front Endocrinol (Lausanne). 2017;8(SEP).

54. Marott SCW, Nordestgaard BG, Tybjaerg-Hansen A, Benn M. Causal associations in type 2 diabetes development. Journal of Clinical Endocrinology and Metabolism. 2019;104(4):1313–1324.

55. Pavone F, Piironen J, Bürkner PC, Vehtari A. Using reference models in variable selection. Comput Stat. 2023;38(1):349–371.

56. IDF Strategic Plan.; 2023.

57. Wang TJ, Larson MG, Vasan RS, et al. Metabolite profiles and the risk of developing diabetes. Nat Med. 2011;17(4):448–453.

58. Keller P, Vollaard NBJ, Gustafsson T, et al. A transcriptional map of the impact of endurance exercise training on skeletal muscle phenotype. J Appl Physiol. 2011;110(1):46–59.

59. Manning AK, Hivert MF, Scott RA, et al. A genome-wide approach accounting for body mass index identifies genetic variants influencing fasting glycemic traits and insulin resistance. Nat Genet. 2012;44(6):659–669.

60. Crossland H, Smith K, Idris I, Phillips BE, Atherton PJ, Wilkinson DJ. Exploring mechanistic links between extracellular branched-chain amino acids and muscle insulin resistance: an in vitro approach. Am J Physiol Cell Physiol. 2020;319(6):C1151–C1157.

61. Roth Flach RJ, Bollinger E, Reyes AR, et al. Small molecule branched-chain ketoacid dehydrogenase kinase (BDK) inhibitors with opposing effects on BDK protein levels. Nat Commun. 2023;14(1).

62. Sitryawan A, Hawes JW, Harris RA, Shimomura Y, Jenkins AE, Hutson SM. A molecular model of human branched-chain amino acid metabolism. Am J Clin Nutr. 1998;68(1):72–81.

63. Shin AC, Fasshauer M, Filatova N, et al. Brain Insulin Lowers Circulating BCAA Levels by Inducing Hepatic BCAA Catabolism. Cell Metab. 2014;20(5):898–909.

64. Sayda MH, Abdul Aziz MH, Gharahdaghi N, et al. Caloric restriction improves glycaemic control without reducing plasma branched-chain amino acids or keto-acids in obese men. Sci Rep. 2022;12(1).

65. Pavone F, Piironen J, Bürkner PC, Vehtari A. Using reference models in variable selection. Comput Stat. 2023;38(1):349–371.

66. Piironen J, Paasiniemi M, Vehtari A. Projective inference in high-dimensional problems: Prediction and feature selection. 10.1214/20-EJS1711. 2020;14(1):2155-2197.

67. Tripathy D, Cobb JE, Gall W, et al. A novel insulin resistance index to monitor changes in insulin sensitivity and glucose tolerance: The ACT NOW study. Journal of Clinical Endocrinology and Metabolism. 2015;100(5):1855–1862.

68. Wallace TM, Levy JC, Matthews DR, Homa T. Use and Abuse of HOMA Modeling. Diabetes Care. 2004;27(6):1487–1495.

69. Levy JC, Matthews DR, Hermans MP. Correct homeostasis model assessment (HOMA) evaluation uses the computer program. Diabetes Care. 1998;21(12):2191–2192.

70. Matsuda M, Defronzo RA. Insulin Sensitivity Indices Obtained From Comparison with the euglycemic insulin clamp. Diabetes Care. 1999;22(9):1462–1470.

71. Utzschneider KM, Prigeon RL, Faulenbach M V., et al. Oral Disposition index predicts the development of future diabetes above and beyond fasting and 2-h glucose levels. Diabetes Care. 2009;32(2):335–341.

