## Supplementary figures and images for "Biomarkers of insulin resistance and their performance as predictors of treatment response in overweight adults"

### Figure S1

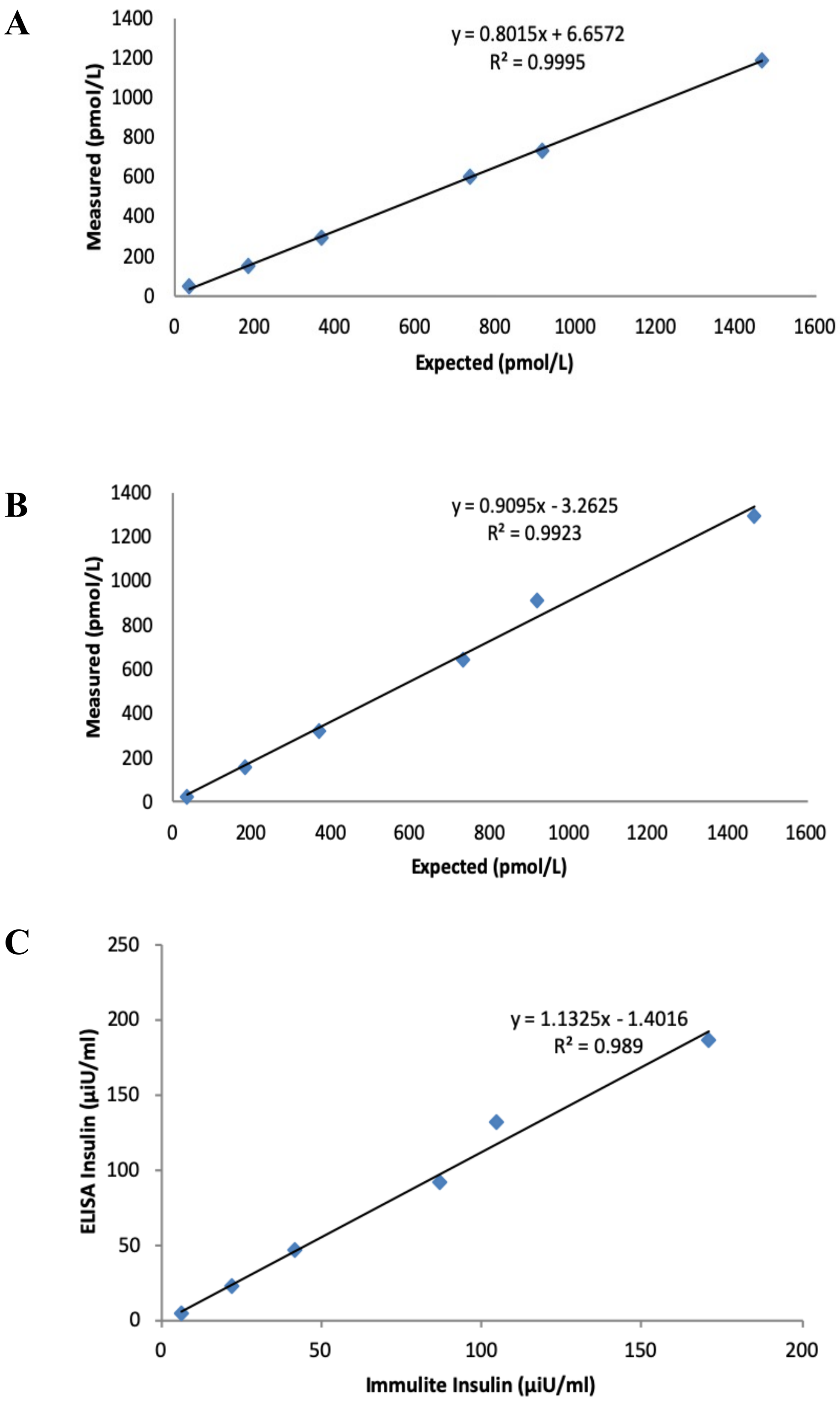

### Figure S2

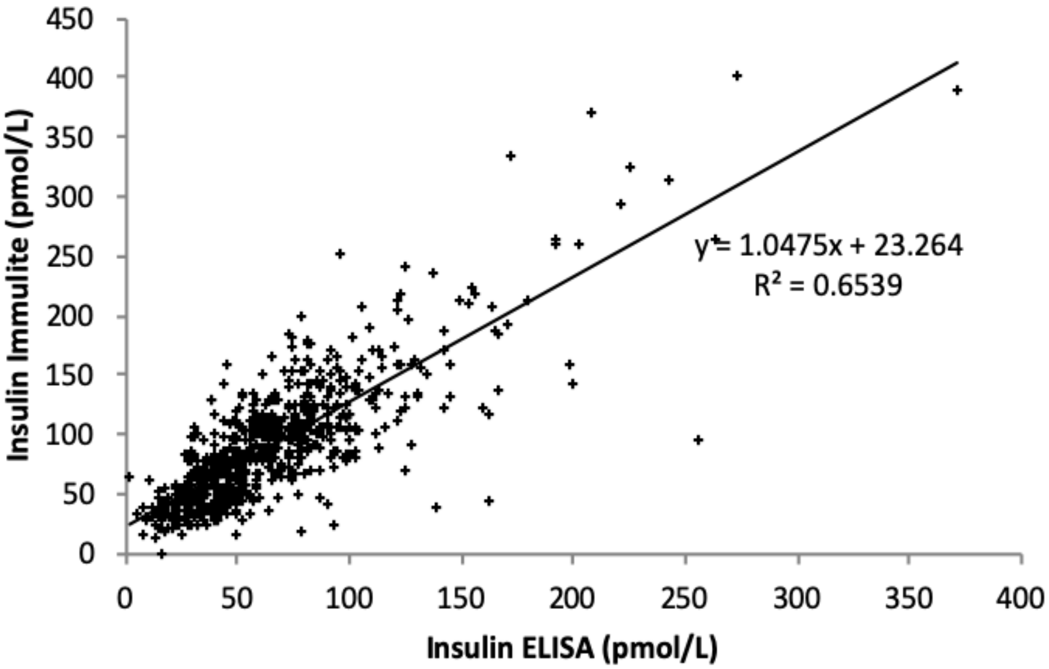

### Figure S3

#### Figure S3

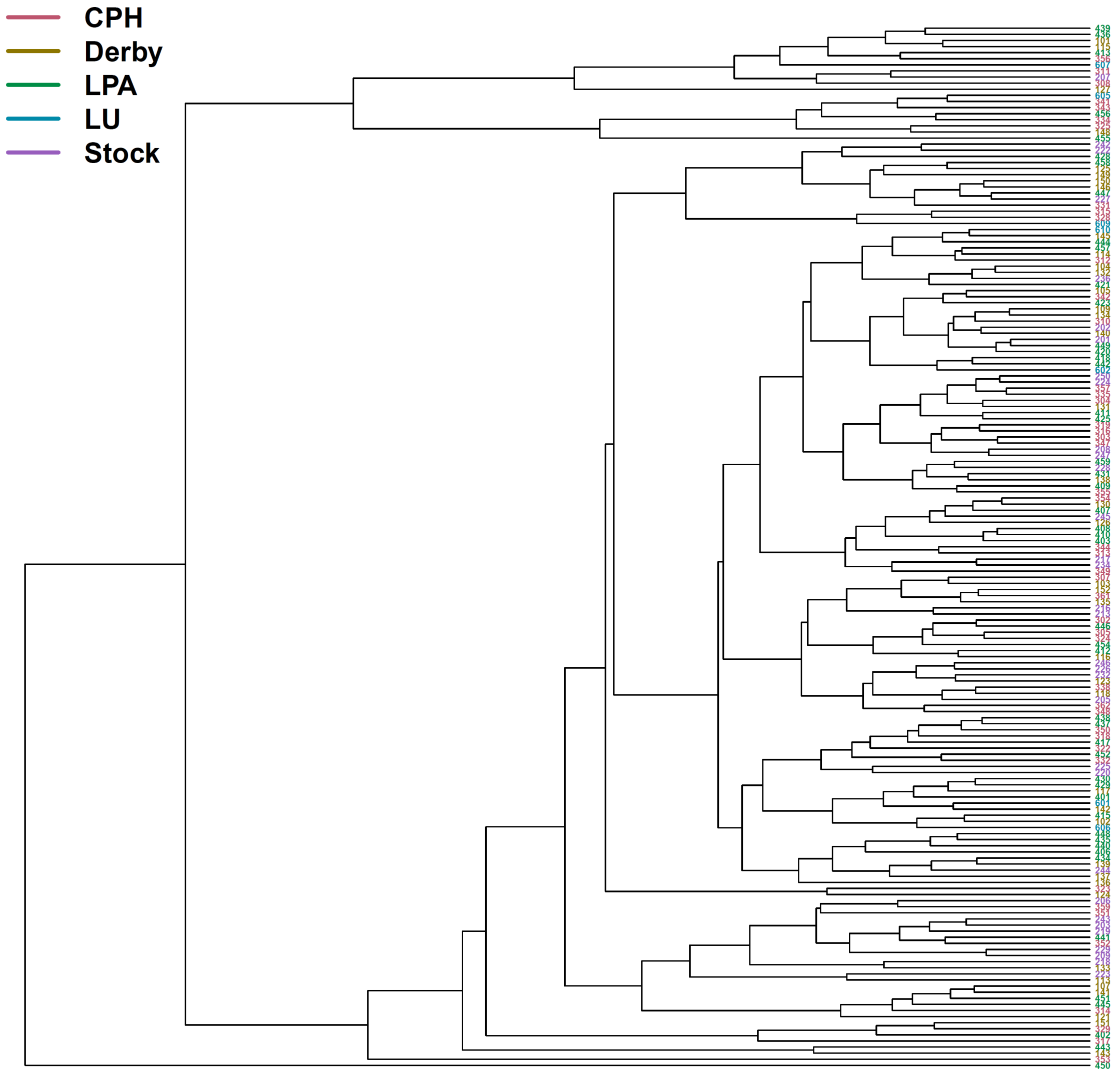

### Figure S4

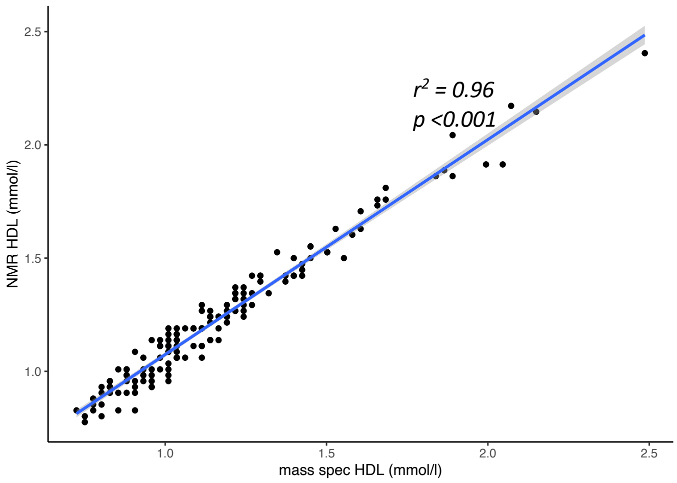

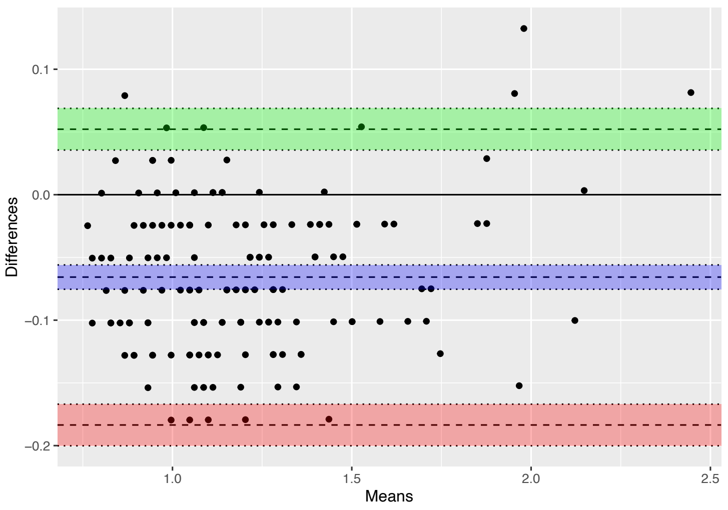

### Figure S5

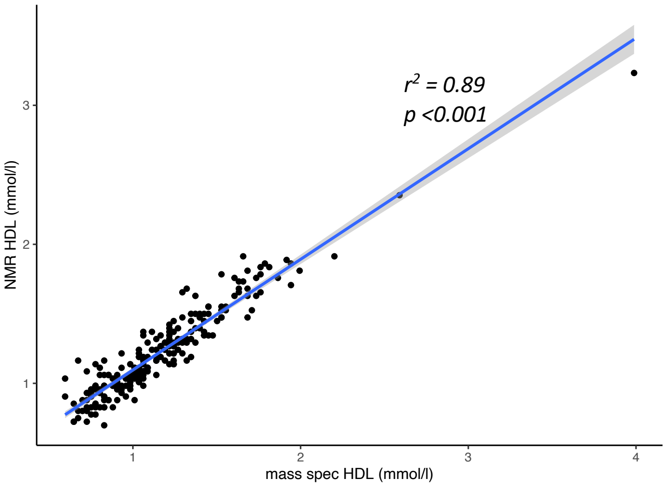

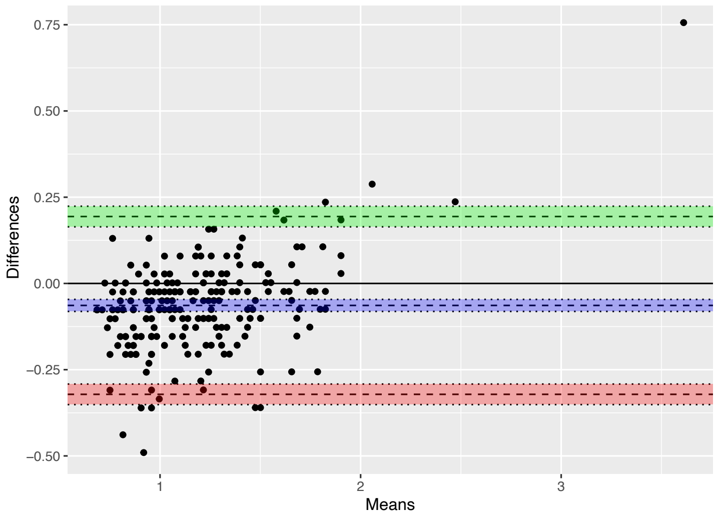

### Figure S6

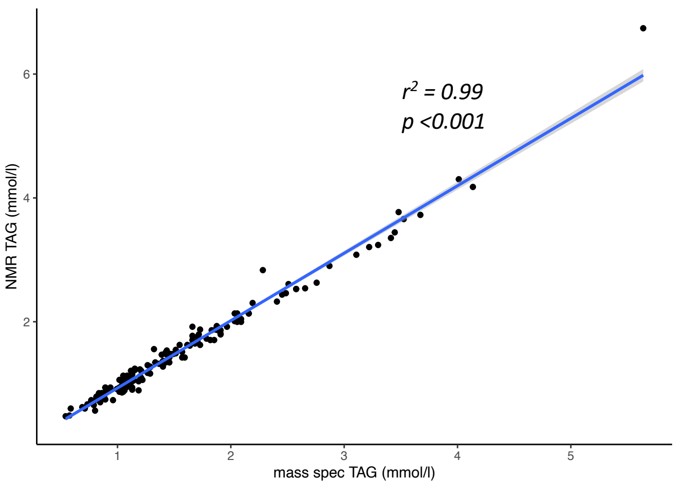

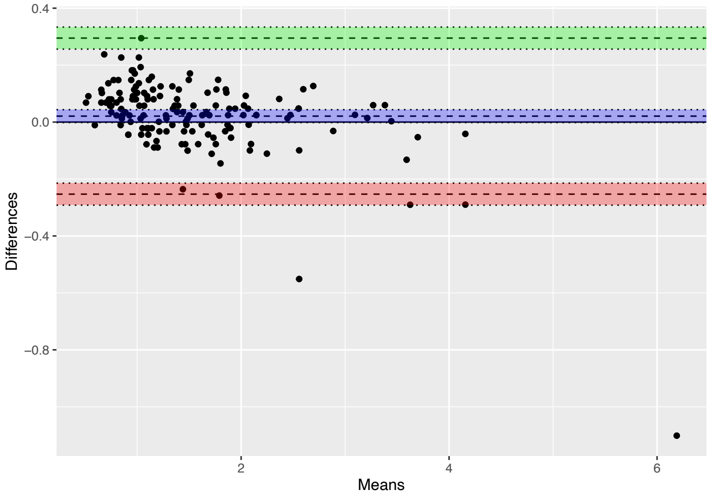

### Figure S7

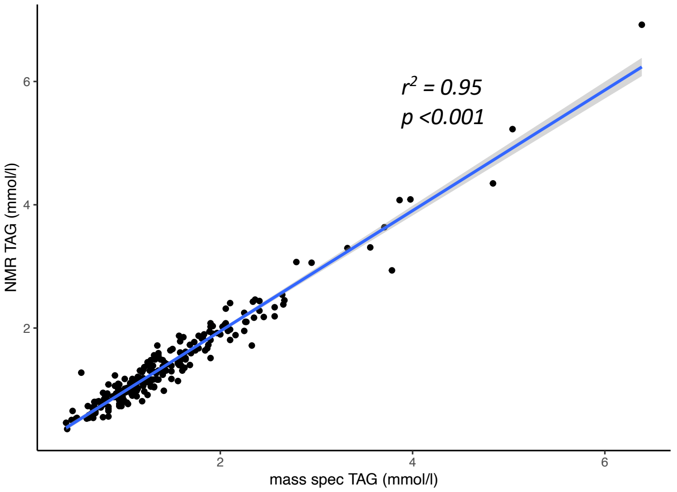

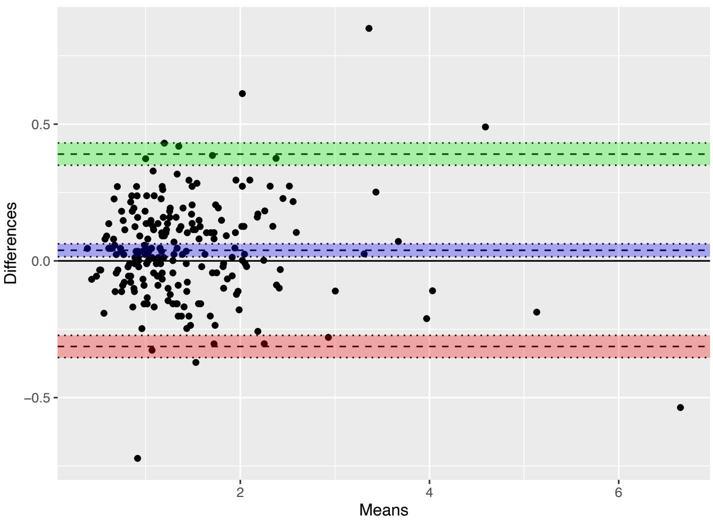

### Figure S8

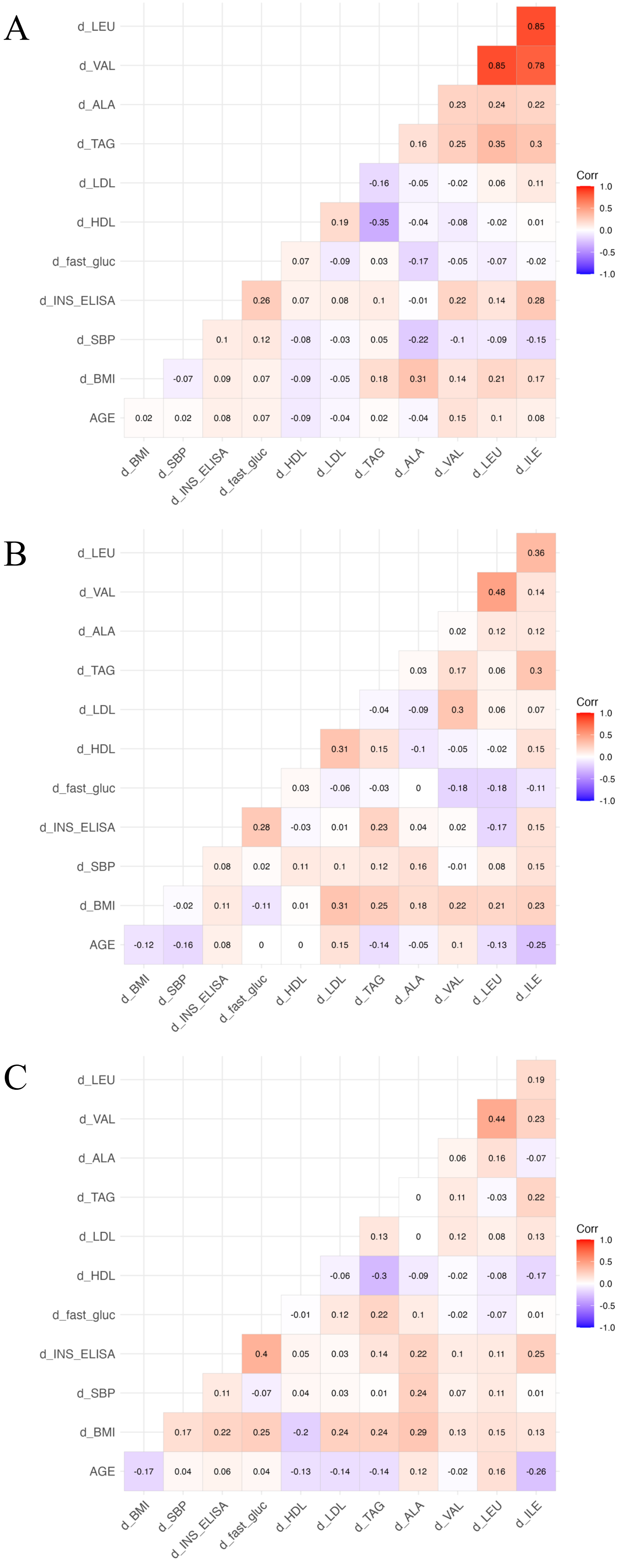
