## Supplementary material for "Biomarkers of insulin resistance and their performance as predictors of treatment response in overweight adults": Table S1

| **META-PREDICT** | | | | | | | |
| --- | --- | --- | --- | --- | --- | --- | --- |
| **Model** | **n** | **Intercept p** | **Intercept estimate** | **Estimates** | **95% CI** | **p** | **Adj r^2^** |
| Age | 188 | <0.0001 | 1.8737 | -0.0032 | -0.0069 – 0.0005 | 0.0918 | 0.010 |
| Gender | 186 | <0.0001 | 1.8254 | -0.1227 | -0.1928 – -0.0527 | **0.0007** | 0.056 |
| BMI (log10) | 188 | 0.5690 | -0.2714 | 1.3532 | 0.7283 – 1.9781 | **<0.0001** | 0.084 |
| SBP | 175 | <0.0001 | 1.1684 | 0.0048 | 0.0016 – 0.0080 | **0.0038** | 0.042 |
| DBP | 174 | <0.0001 | 1.2888 | 0.0060 | 0.0020 – 0.0099 | **0.0030** | 0.044 |
| MAP | 174 | <0.0001 | 1.1370 | 0.0066 | 0.0026 – 0.0106 | **0.0013** | 0.053 |
| Fasting glucose | 188 | 0.1037 | -51.5267 | 25.7240 | 12.2694 – 39.1787 | **0.0002** | 0.066 |
| HDL | 185 | <0.0001 | 2.0214 | -0.2481 | -0.3604 – -0.1358 | **<0.0001** | 0.089 |
| LDL | 184 | <0.0001 | 1.5896 | 0.0599 | 0.0157 – 0.1041 | **0.0082** | 0.032 |
| Triglycerides | 187 | <0.0001 | 1.6140 | 0.1110 | 0.0615 – 0.1604 | **<0.0001** | 0.091 |
| Sum of BCAA | 184 | <0.0001 | 1.3981 | 0.0008 | 0.0005 – 0.0011 | **<0.0001** | 0.123 |
| Isoleucine | 184 | <0.0001 | 1.4579 | 0.0045 | 0.0027 – 0.0063 | **<0.0001** | 0.115 |
| Leucine | 184 | <0.0001 | 1.4408 | 0.0025 | 0.0015 – 0.0036 | **<0.0001** | 0.106 |
| Valine | 184 | <0.0001 | 1.3818 | 0.0015 | 0.0009 – 0.0021 | **<0.0001** | 0.126 |
| Alanine | 184 | <0.0001 | 1.5930 | 0.0005 | 0.0001 – 0.0009 | **0.0148** | 0.027 |
| **STRRIDE-2** | | | | | | | |
| **Model** | **n** | **Intercept p** | **Intercept estimate** | **Estimates** | **95% CI** | **p** | **Adj r^2^** |
| Age | 128 | <0.0001 | 1.7792 | -0.0027 | -0.0082 – 0.0027 | 0.3227 | 0 |
| Gender | 128 | <0.0001 | 1.7628 | -0.0886 | -0.2063 – 0.0290 | 0.1386 | 0.009 |
| BMI (log10) | 128 | 0.0024 | -2.8007 | 3.0012 | 1.7913 – 4.2110 | **<0.0001** | 0.154 |
| SBP | 111 | <0.0001 | 1.2972 | 0.0029 | -0.0017 – 0.0076 | 0.2139 | 0.005 |
| DBP | 111 | 0.0001 | 1.1314 | 0.0066 | -0.0004 – 0.0136 | 0.0650 | 0.022 |
| MAP | 111 | 0.0002 | 1.1428 | 0.0055 | -0.0009 – 0.0119 | 0.0911 | 0.017 |
| Fasting glucose | 118 | 0.0001 | 0.9260 | 0.1394 | 0.0582 – 0.2206 | **0.0009** | 0.083 |
| HDL | 128 | **<0.0001** | 1.8172 | -0.2448 | -0.4208 – -0.0687 | **0.0068** | 0.049 |
| LDL | 128 | **<0.0001** | 1.8172 | -0.0606 | -0.1486 – 0.0274 | 0.1753 | 0.007 |
| Triglycerides | 128 | **<0.0001** | 1.4320 | 0.1329 | 0.0677 – 0.1981 | **0.0001** | 0.107 |
| Sum of BCAA | 126 | **<0.0001** | 0.8578 | 0.0017 | 0.0009 – 0.0024 | **0.0001** | 0.117 |
| Isoleucine | 126 | **<0.0001** | 1.2102 | 0.0066 | 0.0029 – 0.0102 | **0.0005** | 0.085 |
| Leucine | 126 | **<0.0001** | 1.1699 | 0.0027 | 0.0008 – 0.0047 | **0.0064** | 0.051 |
| Valine | 126 | **<0.0001** | 0.8261 | 0.0035 | 0.0019 – 0.0050 | **<0.0001** | 0.13 |
| Alanine | 128 | **<0.0001** | 1.2359 | 0.0010 | 0.0003 – 0.0016 | **0.0027** | 0.062 |
| **STRRIDE PD** | | | | | | | |
| **Model** | **n** | **Intercept p** | **Intercept estimate** | **Estimates** | **95% CI** | **p** | **Adj r^2^** |
| Age | 155 | <0.0001 | 1.9753 | -0.0046 | -0.0116 – 0.0024 | 0.1944 | 0.005 |
| Gender | 155 | <0.0001 | 1.7253 | -0.0395 | -0.1455 – 0.0665 | 0.463 | 0 |
| BMI (log10) | 155 | 0.1983 | -1.2123 | 1.9683 | 0.7168 – 3.2198 | **0.0023** | 0.053 |
| SBP | 134 | <0.0001 | 1.2980 | 0.0032 | -0.0009 – 0.0074 | 0.1260 | 0.010 |
| DPB | 134 | <0.0001 | 1.4408 | 0.0035 | -0.0025 – 0.0096 | 0.2507 | 0.002 |
| MAP | 134 | <0.0001 | 1.2897 | 0.0045 | -0.0014 – 0.0105 | 0.1365 | 0.009 |
| Fasting glucose | 154 | 0.2343 | 0.3187 | 0.2357 | 0.1464 – 0.3251 | **<0.0001** | 0.146 |
| HDL | 153 | <0.0001 | 2.0821 | -0.3326 | -0.4602 – -0.2051 | **<0.0001** | 0.144 |
| LDL | 151 | <0.0001 | 1.5234 | 0.0568 | -0.0180 – 0.1316 | 0.1355 | 0.008 |
| Triglycerides | 153 | <0.0001 | 1.5573 | 0.0961 | 0.0311 – 0.1610 | **0.0040** | 0.047 |
| Sum of BCAA | 155 | <0.0001 | 1.1048 | 0.0014 | 0.0007 – 0.0021 | **0.0001** | 0.091 |
| Isoleucine | 155 | <0.0001 | 1.2519 | 0.0071 | 0.0038 – 0.0104 | **<0.0001** | 0.098 |
| Leucine | 155 | <0.0001 | 1.3715 | 0.0022 | 0.0004 – 0.0041 | **0.0159** | 0.031 |
| Valine | 155 | <0.0001 | 0.9665 | 0.0033 | 0.0021 – 0.0046 | **<0.0001** | 0.147 |
| Alanine | 155 | <0.0001 | 1.4536 | 0.0006 | -0.0000 – 0.0013 | 0.0675 | 0.015 |
