## Supplementary material for "Biomarkers of insulin resistance and their performance as predictors of treatment response in overweight adults": Table S2

| **Cohort** | **Model** | **RMSE** | **MAE** | **r^2^** | **Adj r^2^** | **p value** |
| --- | --- | --- | --- | --- | --- | --- |
| **MP**  n=179 | 1 | 0.21 | 0.17 | 0.27 | 0.26 | <0.001 |
|  | 2 | 0.21 | 0.16 | 0.3 | 0.28 | <0.001 |
|  | 3 | 0.21 | 0.16 | 0.31 | 0.28 | <0.001 |
|  | 4 | 0.21 | 0.16 | 0.33 | 0.3 | <0.001 |
| **S-2**  n=116 | 1 | 0.27 | 0.22 | 0.27 | 0.24 | <0.001 |
|  | 2 | 0.27 | 0.22 | 0.3 | 0.27 | <0.001 |
|  | 3 | 0.26 | 0.1 | 0.37 | 0.33 | <0.001 |
|  | 4 | 0.25 | 0.21 | 0.39 | 0.34 | <0.001 |
| **S-PD**  n=149 | 1 | 0.3 | 0.24 | 0.2 | 0.18 | <0.001 |
|  | 2 | 0.28 | 0.23 | 0.27 | 0.24 | <0.001 |
|  | 3 | 0.29 | 0.23 | 0.3 | 0.27 | <0.001 |
|  | 4 | 0.29 | 0.22 | 0.34 | 0.3 | <0.001 |
| Model 1 = age+gender+log_10_BMI+fasting glucose  Model 2 = model 1 + sumBCAA  Model 3 = model 1 + HDL+LDL+TAG  Model 4 = model 1 + HDL+LDL+TAG+sumBCAA | | | | | | |
